# Optimizing Family Self-Efficacy Through Self-Care Guidelines To Reduce Rehospitalization Rates In Stroke Patients: A Quasi-experimental study

**DOI:** 10.1101/2024.12.20.24319475

**Authors:** Komang Ardidhana Nugraha Putra, Moses Glorino Rumambo Pandin, Ni Luh Putu Ayu Ratna Dewi, Alfi Shari

## Abstract

**Introduction:** Since 2007, there has been an alarming spike in stroke cases in Indonesia, leading to an increase in rehospitalization due to complications and recurrent stroke attacks. Self-care with family support has been proven effective in supporting stroke recovery.

**Objective:** This study aims to assess the effectiveness of self-care guidelines to prevent rehospitalization of stroke patients on family self-efficacy.

**Method:** A quasi-experimental research design was used in this study. A total of 80 respondents were involved, consisting of 40 respondents in each group. Data collection was carried out using observation sheets and the General Self-Efficacy Scale (GSE).

**Results:** There was an increase in self-efficacy in the intervention group before and after the implementation of self-care guidelines in families caring for stroke patients with a p value <0.001.

**Conclusion:** The implementation of self-care guidelines is effective in increasing family self-efficacy in long-term care to prevent rehospitalization of stroke patients. Evaluation of the long-term impact of implementing guidelines on rehospitalization and quality of life of stroke patients is important to do.

## INTRODUCTION

Stroke is the second leading cause of death and the third leading cause of disability in the world (American Stroke Association, 2019). The prevalence of stroke in the world continues to increase every year. In 2019, there were 12.2 million new cases of stroke and 101 million people living with stroke in the world (Feigin et al., 2021). Of the total cases in the world, Asia is the highest contributor of cases, namely 58.1 million cases. Indonesia is the country with the second highest stroke rate after Mongolia, namely 193.3 per 100,000 person-years (Venketasubramanian et al., 2017). Indonesia experienced an increase in stroke cases of 2.1‰ in five years based on data from the 2013 and 2018 Riskesdas. East Kalimantan is the province with the highest number of stroke cases, namely 14.7‰ and Bali Province is ranked 17th with a stroke case rate of 10.7‰ based on data from the 2018 Riskesdas. Judging from the increase in cases, Bali Province experienced a significant number of cases in a period of 5 years, namely a two-fold increase based on data from the 2013 and 2018 Riskesdas (Riskesdas, 2018).

Stroke survivors who survive their first stroke are also at risk of recurrence. The American Stroke Association (2022) states that one in four patients is at risk of stroke recurrence. Based on the research results of Trisetiawati et al. (2018) found that 45.8% of 238 stroke patients had experienced recurrent strokes. Research conducted by Flach, Muruet, Wolfe, Bhalla, and Douiri (2020) found an increase in recurrence cases of 35% (1995-1999) and 67% (2010-2015). One of the causes of the high number of recurrences is the lack of awareness of stroke patients to carry out routine checks at health facilities. Research by Trisetiawati et al. (2018) found that stroke patients who did not control their condition regularly after the first stroke had an 8.7 times higher risk of stroke recurrence than the first stroke.

Stroke sufferers to carry out routine checks at health facilities in Indonesia nationally are still very minimal, namely 39.4%. Meanwhile, in Bali Province, the awareness of stroke sufferers to carry out control is higher than the national average, but still less than half of sufferers carry out routine control, namely 44% (Riskesdas, 2018). According to the World Health Organization (2021), one way to better control stroke is to involve patients in monitoring their own self-care. Many studies related to self-care have been conducted and have been proven effective in improving quality of life and self-efficacy (Fryer et al., 2016). Resulting in improved conditions, as well as recovery from adverse effects such as dependence and death (Parke et al., 2015).

Research related to the development of self-care guidelines to prevent rehospitalization of stroke patients has been conducted using the Delphi study method (Putra et al., 2024). However, the effectiveness of implementing self-care guidelines in improving the self-efficacy of families caring for stroke patients still needs to be studied further. Studies that strengthen the evidence regarding the effectiveness of self-care guidelines can help families improve post-hospital care for stroke patients, reduce the burden of rehospitalization, and improve the quality of life of patients and their families. Thus, research on the effectiveness of implementing self-care guidelines on family self-efficacy in caring for and preventing rehospitalization of stroke patients has significant clinical and social relevance. This can provide a basis for the development and implementation of more effective interventions in the management of stroke patients, thereby improving long-term health outcomes for patients and optimizing the use of health resources.

## METHODS

### Research design

This study used a quasi-experimental two groups pre and post-test approach. The quasi-experimental approach is a type of experimental research that provides manipulation of independent variables, but without randomization in the selection between treatment groups and control groups (Swarjana, 2015). This study was conducted to determine the effectiveness of self-care guidelines in improving the self-efficacy of families caring for stroke patients.

### Population, samples, and Sampling

The population of this study was families who actively care for stroke patients. The final sample size in each group was 40 respondents with a total sample size of 80 respondents. The sample calculation formula used for this experimental study was the Rosner sample size formula. The inclusion and exclusion criteria in this study were families who actively care for stroke patients for > 3 months, families with good cognitive and physical adequacy, and families who were willing to be respondents by signing an informed consent.

### Instruments and Data Collection

Data collection in this study used observation sheets and the General Self-Efficacy Scale (GSE) questionnaire. The GSE questionnaire is an evaluation tool used to measure the general beliefs of families caring for stroke patients in their own abilities to overcome challenges, deal with problems, and achieve goals in everyday life.

### Data Aalysis

The data analysis used included univariate and bivariate analysis, which began with an assumption test because the scales used were nominal (Self-care guidelines) and interval (self-efficacy). The normality test was carried out using Shapiro-Wilk on the characteristics of the respondents (age, gender, education, marital status, occupation), and the homogeneity test with chi-square. The univariate analysis aimed to identify the general characteristics of the respondents (age, gender, marital status, education level, occupation), the level of self-efficacy before and after being given self-screening guidelines in the intervention and control groups, and prevention of rehospitalization in stroke patients. The data were presented in a frequency distribution table that included the mean, standard deviation, maximum value, and minimum value. Bivariate analysis used the Paired T-Test or Wilcoxon Signed-Rank Test to test the differences in the effectiveness of self-screening guidelines in each group, followed by the Independent T-Test to compare the differences between the intervention and control groups. This analysis also tested the effect of self-screening guidelines on self-efficacy and rehospitalization in both groups, as well as a comparison of their effects. All data were analyzed using SPSS 24.

**Figure 1.**
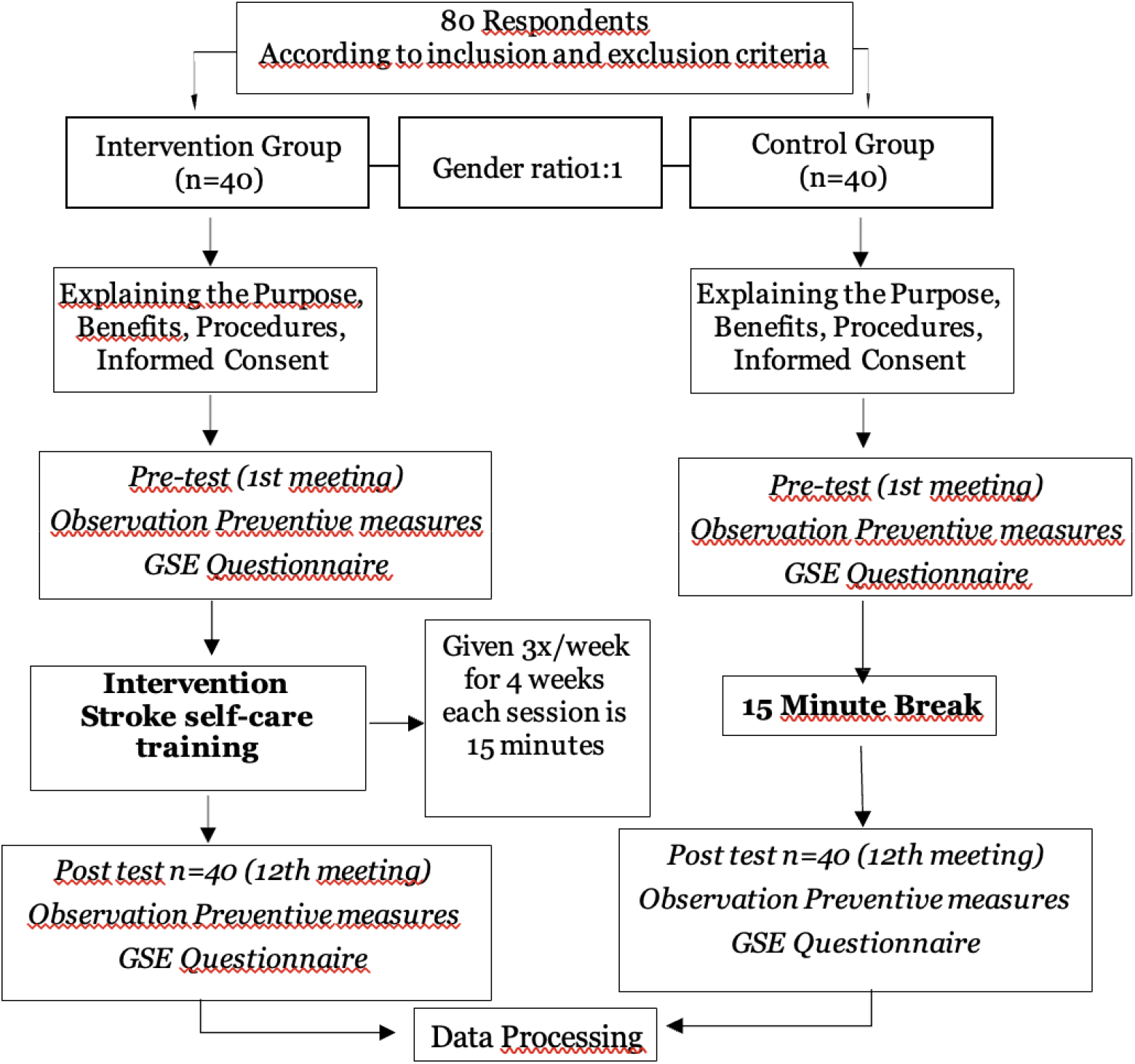
Operational research framework

### Ethical Consideration

This study received ethical approval from the Research Ethics Commission of the Faculty of Health, Bali Institute of Technology and Health (ethical license number 04.0529/KEPITEKES- BALI/X/2023). All participants gave informed consent and were made aware of their right to disengage from the study without negative consequences. This research applies the ethical principle of anonymity to protect the identity and confidentiality of expert panel information by using codes or initials, deleting personal identification information, and monitoring data security.

## RESULT

The study was conducted in the working area of the Bangli District Health Office. The general characteristics of the respondents can be seen in table 1.

**Table 1.** Respondent characteristics.

| Characteristics | <u>Intervention</u> |  | <u>Control</u> |  |
| --- | --- | --- | --- | --- |
|  | f | % | f | % |
| **Age (years)** |  |  |  |  |
| 20-30 | 5 | 12,5 | 4 | 10,0 |
| 31-40 | 10 | 25,0 | 12 | 30,0 |
| 41-50 | 15 | 37,5 | 14 | 35,0 |
| 51-60 | 8 | 20,0 | 7 | 17,5 |
| >60 | 2 | 5,0 | 3 | 7,5 |
| **Gender** |  |  |  |  |
| Manle | 20 | 50,0 | 20 | 50,0 |
| Female | 20 | 50,0 | 20 | 50,0 |
| **Relationship with patients** |  |  |  |  |
| Husband/Wife | 10 | 25,0 | 8 | 20,0 |
| Children | 27 | 67,5 | 29 | 72,5 |
| Other Family | 3 | 7,5 | 3 | 7,5 |
| **Level of education** |  |  |  |  |
| Elementary School | 2 | 5,0 | 3 | 7,5 |
| Junior High School | 5 | 12,5 | 6 | 15,0 |
| High School | 14 | 35,0 | 15 | 37,5 |
| College | 19 | 47,5 | 16 | 40,0 |

Based on table 1 shows that the age range of most respondents is between 41-50 years with 15 (37.5%) in the intervention group and 14 (35%) in the control group. Based on gender in the intervention group has a ratio of 1:1 in each group. Characteristics based on relationships with patients are mostly child relationships with 27 (67.5%) in the intervention group and 29 (72.5%) in the control group. The level of education is mostly at the college level with a total of 19 (47.5%) in the intervention group and 16 (40.0%) in the control group.

The results of self-efficacy before and after the implementation of self-care guidelines in the intervention and control groups were categorized into four, namely very low, low, moderate, and high. Furthermore, the anxiety score was tested for normality using Shapiro-Wilk and homogeneity test using Lavene statistics which can be seen in Table 2.

**Table 2.** Family self-efficacy results before and after implementing self-care guidelines.

| Characteristics | <u>Intervention</u> |  |  | <u>Control</u> |  |  | Homogeneity<br>(*P-Value*) |
| --- | --- | --- | --- | --- | --- | --- | --- |
|  | f (%) | Mean | SD | f (%) | Mean | SD |  |
| *Pre-test* |  |  |  |  |  |  |  |
| Very low | 10 (25%) | 30,5 | 10,2 | 12 (30%) | 29,0 | 9,5 | 0,356** |
| Low | 15 (37,5%) |  |  | 14 (35%) |  |  |  |
| Medium | 10 (25%) |  |  | 9 (22,5%) |  |  |  |
| High | 5 (12,5%) |  |  | 5 (12,5%) |  |  |  |
| Normality (P-Value) | 0,128* |  |  | 0,200* |  |  |  |
| *Post-test* |  |  |  |  |  |  |  |
| Very low | 2 (5%) | 40.2 | 8.5 | 11 (27,5%) | 28,5 | 10,0 | 0,294** |
| Low | 5 (12,5%) |  |  | 15 (37,5%) |  |  |  |
| Medium | 9 (22,5%) |  |  | 10 (25%) |  |  |  |
| High | 24 (60%) |  |  | 4 (10%) |  |  |  |
| Normality (P-Value) | 0,076* |  |  | 0,157* |  |  |  |

\* Shapiro-wilk
\*\* Lavenne Statistic

Based on table 2, the results of the pre-test self-efficacy in the intervention group were mostly at a low level of 15 (37.5%), while the results of the post-test showed that most respondents experienced an increase in the high category of 24 (60%). In the control group, the pre-test results were mostly at a low self-efficacy level of 14 (35%), while the results of the post-test, most respondents remained at a low self-efficacy level of 15 (37.5%). When viewed from the mean value, an increase in the level of self-efficacy occurred in the intervention group and conversely there was a decrease in the average self-efficacy score in the control group. The normality test using Shapiro-Wilk obtained normally distributed data on the pre-test and post-test results of the intervention group and the control group with a p-value> 0.05. The results of the homogeneity test with the Lavene statistic obtained pre-test and post-test results with a p-value of 0.356> 0.05 and 0.294> 0.05 so that the data was declared homogeneous.

Analysis of differences in family self-efficacy using paired t-tests was conducted on each intervention group and control group, which can be seen in Table 3.

**Table 3.** Analysis of differences in family self-efficacy.

| Group | <u>Mean±SD</u> |  | <u>Mean±SD</u><br><u>Difference</u> | <u>P-Value</u> |
| --- | --- | --- | --- | --- |
|  | *Pre-test* | *Post-test* |  |  |
| Intervention | 30,5 ± 10,2 | 40.2 ± 8.5 | 9,7 ± 13.27 | <0,001 |
| Control | 29,0 ± 9,5 | 28,5 ± 10,0 | -0,5 ± 13,80 | 0,237 |

Based on table 3, it shows that in the intervention group, a p-value <0.001 was obtained, which means that there was a significant difference in self-efficacy scores in families caring for stroke patients before and after the implementation of self-care guidelines with an average increase of 9.7. While in the control group, a p-value of 0.237> 0.05 was obtained, which means that there was no difference in self-efficacy scores in families before and after the implementation of self-care guidelines with an average decrease of 0.5.

## DISCUSSION

The self-care guidelines applied to families caring for stroke patients in this study were designed to provide practical information and strategies to families in providing care for stroke patients. The points discussed in this guide include self-care activities, including drug therapy, physical exercise, diet and nutrition, stress management, self-motivation, functional status examinations, and risk factor management (Putra et al., 2024). With clear and structured guidelines, families feel more prepared and confident in facing the challenges of caring for stroke patients (Li, M et al., 2023).

The results of this study indicate that families who implement care guidelines in caring for stroke patients experience a significant increase in self-efficacy. In post-stroke care, family knowledge and self-efficacy are very important. Higher family knowledge and self-efficacy are correlated with better family skills in performing rehabilitation exercises, highlighting the importance of educating families to improve patient outcomes (Sari et al., 2023). For chronic conditions such as Stroke, Alzheimer’s and pulmonary TB, family knowledge and self-efficacy are essential for effective care. Families with better knowledge and self-efficacy tend to be able to manage symptoms effectively and ensure patient adherence to treatment (Rachmawati et al., 2022).

Increasing family self-efficacy can contribute directly to preventing rehospitalization of stroke patients. Family resilience, which includes the ability to adapt and support patients, is a predictor of better outcomes in post-stroke recovery. Improving family resilience through increased self-efficacy and social support can lead to better family functioning and reduce the likelihood of rehospitalization in stroke patients (Zhang, 2022).

### Philosophical Studies

In the context of this study, ontology is related to the assumption of existing reality, namely that family self-efficacy is a real and measurable factor that influences stroke patient care. This study also considers the family as a vital care unit in supporting the recovery process and preventing patient rehospitalization. In addition, self-care guidelines are considered an effective tool to increase family capacity in caring for stroke patients independently, with the hope of improving the quality of care and patient outcomes.

Based on epistemological studies, this study is related to how researchers gain knowledge about the influence of self-care guidelines on family self-efficacy. A quasi-experimental approach is used to empirically test hypotheses, relying on structured data collection. The use of measuring instruments such as the General Self-Efficacy Scale (GSE) shows an effort to gain measurable and objective knowledge about self-efficacy. However, methodological limitations, such as the lack of randomization in group selection, can affect the validity of the results and reflect the challenges in gaining knowledge that is completely objective and universally applicable.

The axiology of this study reflects a commitment to humanitarian values and research ethics. The focus of the study on improving the quality of life of stroke patients and their families demonstrates a strong orientation towards human welfare. The researchers also ensured that the study was conducted with respect for ethics, including obtaining ethical approval involving the provision of clear information and informed consent from participants. In addition, the social implications of this study are significant, as the results may contribute to the development of more effective interventions in the care of stroke patients, which in turn may improve the quality of life of patients and have a positive impact on society and the wider health system.

## CONCLUSIONS

This study shows that the implementation of self-care guidelines significantly increases family self-efficacy in caring for stroke patients. By providing clear information and practical steps, these guidelines help families feel more confident and skilled in managing care at home. This increase in self-efficacy has positive implications for the family’s ability to prevent patient rehospitalization. When families have higher confidence in their abilities, they are better able to carry out necessary interventions, and handle changes in the patient’s condition more quickly and effectively. Therefore, the implementation of self-care guidelines is not only beneficial for improving family knowledge and skills, but also contributes to improving the quality of life of stroke patients and reducing the risk of rehospitalization. This study recommends the use of similar guidelines in health care practices and the need for ongoing support for patient families.

## Data Availability

All data produced in the present study are available upon reasonable request to the authors

## Acknowledgement

We thank all participants, family and data collectors for their contribution to this study.

## Notes

### Competing Interest Statement

The authors have declared no competing interest.

### Funding Statement

This study did not receive any funding

### Author Declarations

Research Ethics Commission of the Faculty of Health, Bali Institute of Technology and Health

